# Are women leaders significantly better at controlling the contagion?

**DOI:** 10.1101/2020.06.06.20124487

**Authors:** Soumik Purkayastha, Maxwell Salvatore, Bhramar Mukherjee

## Abstract

Recent media articles have suggested that women-led countries are doing better in terms of their responses to the COVID-19 pandemic. We examine an ensemble of public health metrics to assess the control of COVID-19 epidemic in women- versus men-led countries worldwide based on data available up to June 3. The median of the distribution of median time-varying effective reproduction number for women- and men-led countries were 0.89 and 1.14 respectively with the 95% two-sample bootstrap-based confidence interval for the difference (women - men) being [- 0.34, 0.02]. In terms of scale of testing, the median percentage of population tested were 3.28% (women), 1.59% (men) [95% CI: (−1.29%, 3.60%)] with test positive rates of 2.69% (women) and 4.94% (men) respectively. It appears that though statistically not significant, countries led by women have an edge over countries led by men in terms of public health metrics for controlling the spread of the COVID-19 pandemic worldwide.

**One Sentence Summary:** We quantitatively compare countries led by women with countries led by men in terms of public health metrics for controlling the spread of the novel coronavirus.

## Introduction

The remarkable success of the chancellor of Germany Angela Merkel, the prime minister of New Zealand Jacinda Ardern, the prime minister of Finland Sanna Marin and the Icelandic prime minister Katrín Jakobsdóttir in controlling the COVID-19 pandemic has received much attention (1). There has been a wave of articles in the media that applaud the COVID-19 response efforts in women-led countries (2) along with the promotion of progressive policies which would reduce severe COVID-19 outcomes (3). In this note, we attempt to quantify the effect of women leaders more broadly across the world in terms of public health and policy relevant measures that have been widely discussed in the last three months for controlling a pandemic. Instead of qualitative comparisons and statements, we assess statistical significance of the hypotheses that performance is indeed different between women and men heads of nations.

## Methods

Since the world is full of data, we use data from the Johns Hopkins COVID-19 data repository (4) to carry out a two-group comparison of countries led by men and women. The list of countries with women leaders was retrieved from Wikipedia (5). We restrict our analysis to countries with at least 100 cumulative reported cases of COVID-19 infections and at least 10 days of reported data as of 3^rd^ June 2020. Of the 159 countries so chosen, 18 have women heads of state (*Table 1*) while the other 141 have men heads of state.

**Table 1.** Summary case-count, death count and testing data for 18 countries led by women.

| **Country** | **Total number of tests per 100,000<sup>1,2</sup>** | **Total number of cases per 100,000<sup>1,2</sup>** | **Pct. (%) of population tested<sup>1,3</sup>** | **Pct. (%) of test positive cases<sup>1,4</sup>** | **Total number of deaths per 100,000<sup>1,2</sup>** |
| --- | --- | --- | --- | --- | --- |
| Bangladesh | 167.45 | 23.25 | 0.17 | 13.89 | 0.45 |
| Belgium | 5681.92 | 494.77 | 5.68 | 8.71 | 82.16 |
| Bolivia | 214.73 | 61.13 | 0.21 | 28.47 | 3.43 |
| Denmark | 8291.13 | 197.3 | 8.29 | 2.38 | 10.01 |
| Estonia | 6085.38 | 138.25 | 6.09 | 2.27 | 5.20 |
| Ethiopia | 84.00 | 0.64 | 0.08 | 0.76 | 0.01 |
| Finland | 3187.53 | 119.62 | 3.19 | 3.75 | 5.79 |
| Germany | 4718.05 | 212.79 | 4.72 | 4.51 | 10.27 |
| Greece | 1635.48 | 27.85 | 1.64 | 1.70 | 1.72 |
| Iceland | 17611.72 | 528.64 | 17.61 | 3.00 | 2.93 |
| Nepal | 200.01 | 2.65 | 0.20 | 1.32 | 0.03 |
| New Zealand | 5634.11 | 23.93 | 5.63 | 0.42 | 0.46 |
| Norway | 4474.95 | 154.63 | 4.47 | 3.46 | 4.37 |
| Serbia | 3372.5 | 164.99 | 3.37 | 4.89 | 3.60 |
| Singapore | 3743.3 | 540.41 | 3.74 | 14.44 | 0.41 |
| Slovakia | 3035.25 | 27.75 | 3.04 | 0.91 | 0.51 |
| Myanmar | 41.89 | 0.38 | 0.04 | 0.90 | NA |
| Taiwan | 300.55 | 1.85 | 0.30 | 0.62 | NA |

<sup>1</sup> As of June 3<sup>rd</sup>, 2020. Some countries release testing data a week after the tests were performed.
<sup>2</sup> Figures reported are per 100,000 of population size.
<sup>3</sup> Calculated as $100 \times (\text{total number of tests}/\text{population size})$ .
<sup>4</sup> Calculated as $100 \times (\text{total number of positive cases}/\text{total number of tests})$ .

We first create a plot of the effective time-varying reproduction number at time t, namely, R(t) (6) for all 159 countries (*Figure 1*). Time zero in the figure is defined as the day when each country crossed at least 50 cases. We construct smoothed trajectories of median R(t) across countries, stratified by gender of head of state. We calculate the mean, median and maximum R(t) over the course of the pandemic (from time zero till June 3rd) for each country based on the computed R(t) trajectories. We also consider the most recent value of R (on June 3) to gauge where we stand now. We present density plots of these summaries stratified by gender of head of state, along with most recent values of country-level effective R (*Figure 2*). We create a forest plot of country-specific medians and associated 95% confidence intervals (CI) of R(t) values over the course of the pandemic (*Figure 3*).

**Fig. 1.**
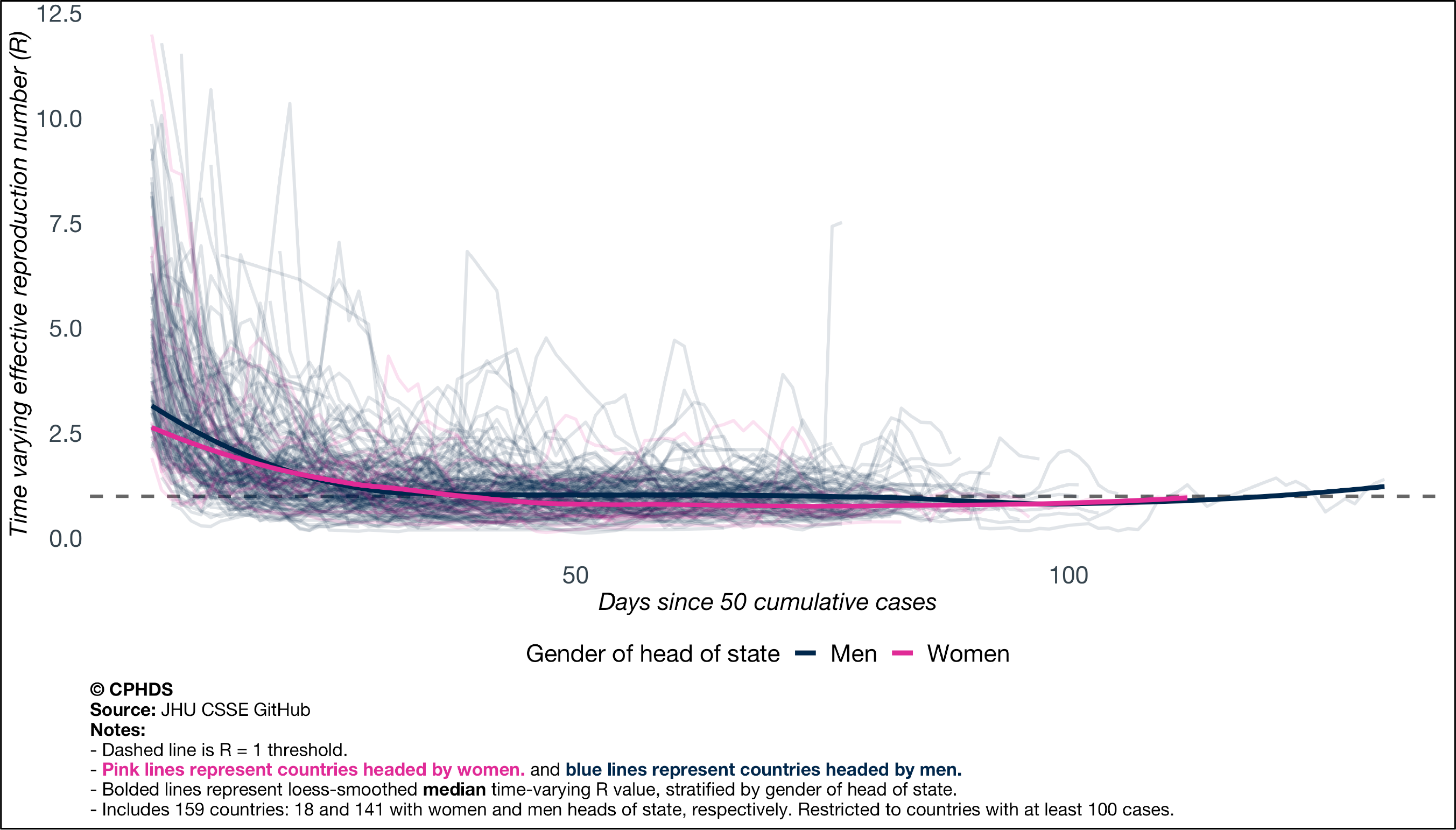
Time varying plot of the effective time varying reproduction number of all 159 countries, with locally smoothed trajectories of median of time-varying reproduction number, stratified by gender of head of state.

**Fig. 2.**
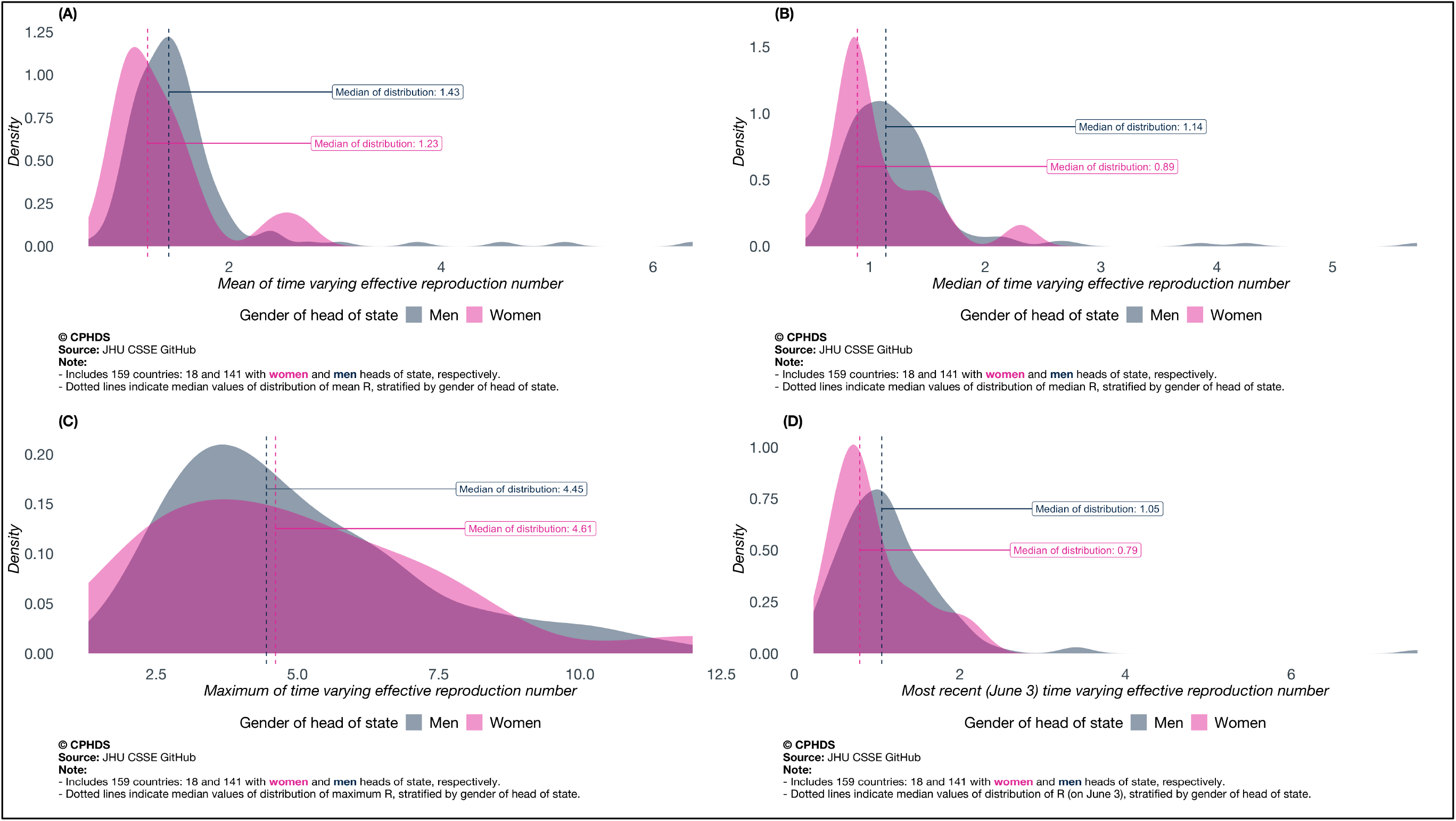
Distributions of (A) mean, (B) median, (C) maxima and (D) most recent values of time-varying reproduction number, stratified by gender of head of state. Mean, median, maximum are taken over the time course with time zero defined as the day the reported counts crossed 50 and the last day considered is June 3.

**Fig. 3.**
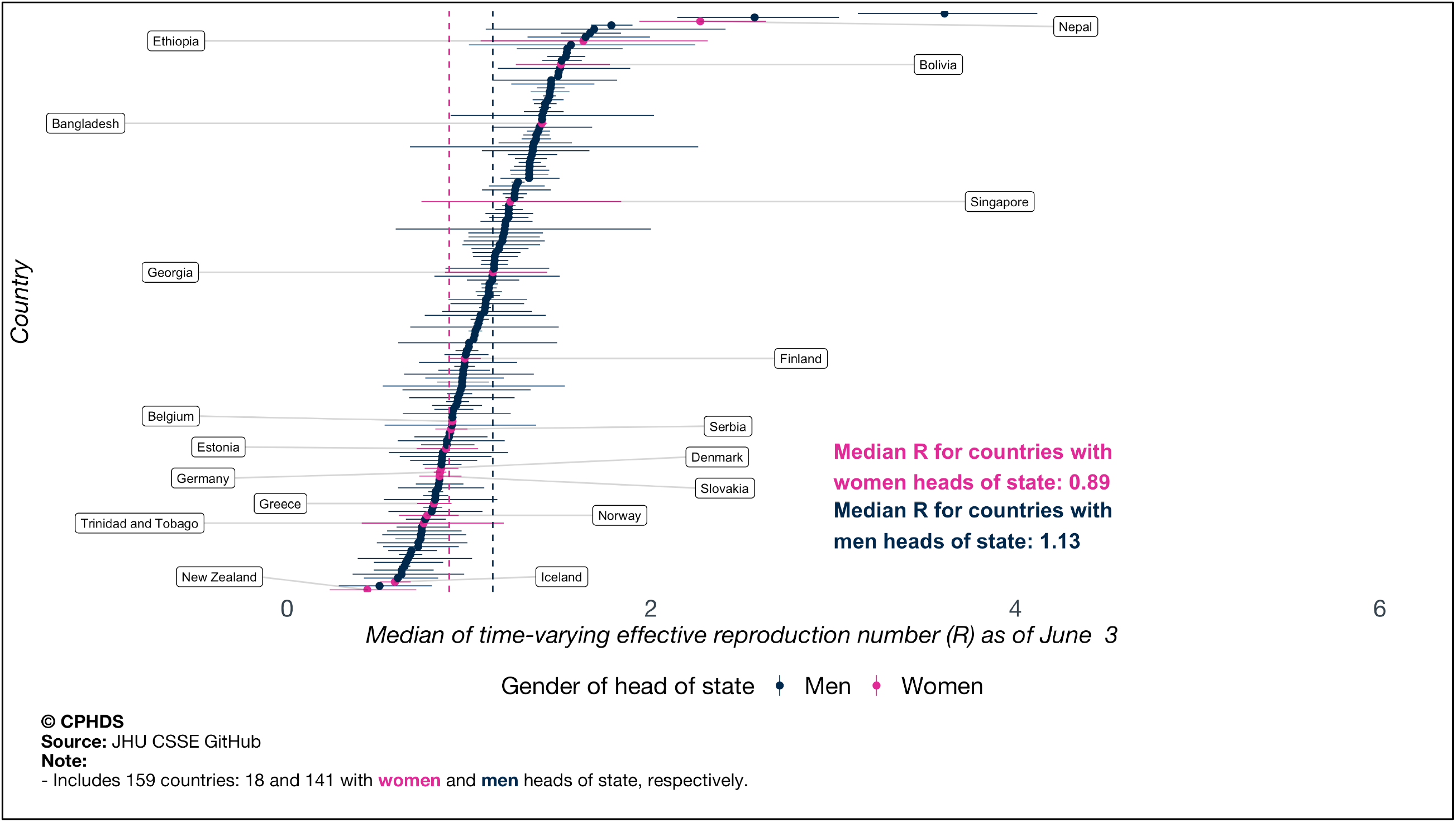
Forest plot of country-specific median and associated 95% CI of time-varying effective reproduction number R, stratified by gender of head of state.

In addition to visual inspection, we do some formal statistical inference. Since the sample sizes of the two groups are unbalanced (18 versus 141) and the distributions are visibly not normal-like, we use resampling procedures. For median, mean, maximum and most recent (as of June 3rd) values of time-varying R(t), we perform a two-sample bootstrap procedure-based comparison of medians and construct 95% CI based on empirical 2.5^th^ and 97.5^th^ percentile of the bootstrap distributions.

We also consider the doubling time (DT) metric and case-fatality rate (CFR) metric under the same analytical lens. To evaluate the scale of diagnostic testing to identify new COVID-19 cases, we compare 85 countries which have released testing data (67 led by men and 18 led by women) on the basis of two metrics: percentage of population tested and percentage of positive tests. We present clustered bar plots comparing percentage of population tested and percentage of positive tests for these 85 countries (*Figure 4*).

**Fig. 4.**
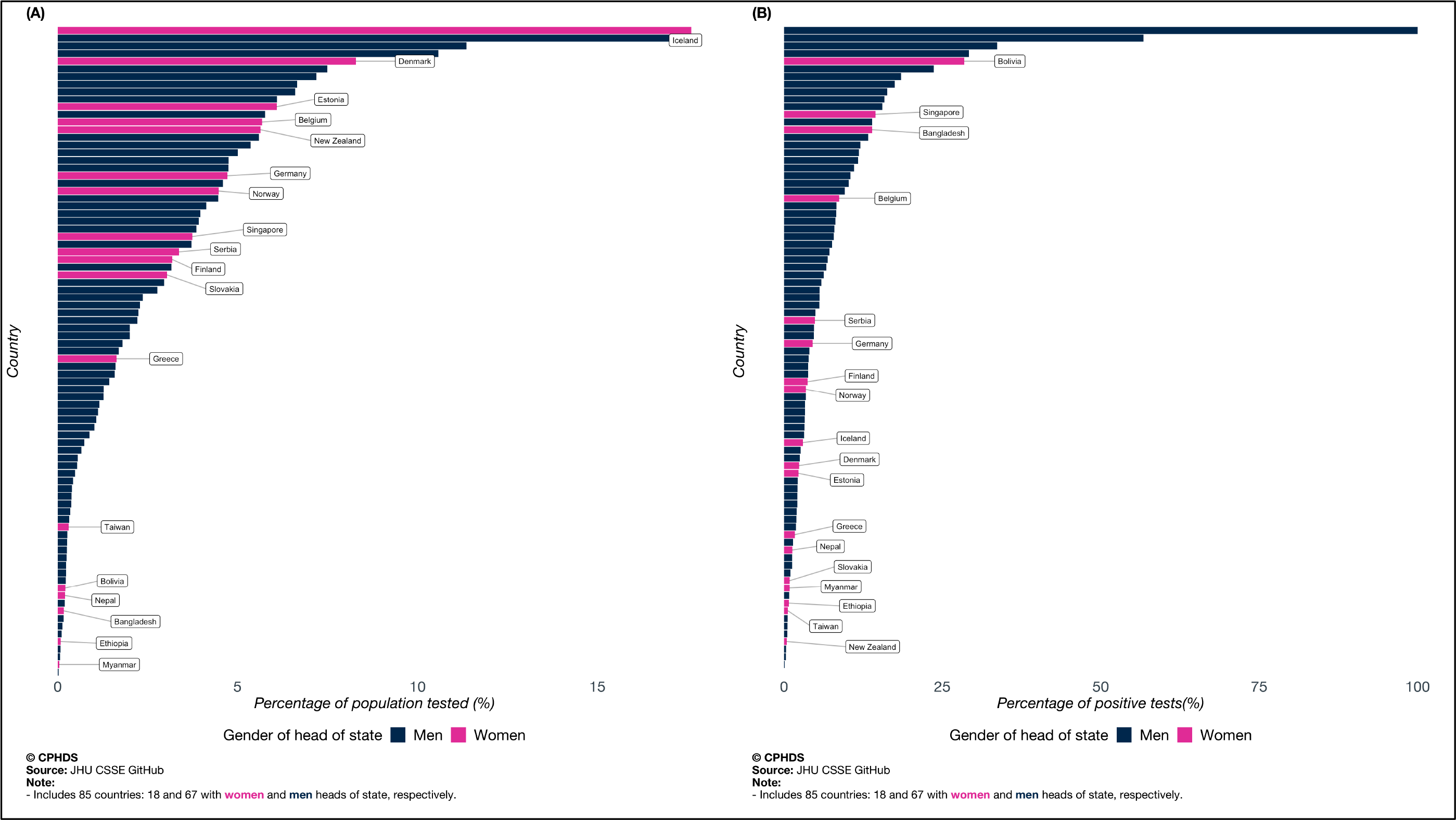
Bar plots of country-specific testing data (A: percentage of population tested and B: percentage of positive tests), stratified by gender of head of state.

## Results

*Table 1* presents a snapshot of the raw data for the 18 countries led by women in terms of numbers of cases and deaths per 100,000 population, percentage of population tested and percentage of test positives. There is significant variation across countries and continents. *Figure 1* presents the median trajectory of the time-varying reproduction number R through the time-course of the pandemic and it shows lower trend for the trajectory for countries led by women. *Figure 2* reveals a shift in the gender-stratified distributions of median, mean, maximum and most recent values of R(t) --- they have more probability weight towards smaller values for the women (lower the R, slower is the transmission). Comparing the two distributions (women vs men) on the basis of median of time-varying R(t) yields a median value of 0.89 for women and 1.14 for men. Values of R *below one* are desirable. The bootstrap procedure comparing difference of median (women-men) yields a 95% CI of [-0.34, 0.02]. Similar comparisons between gender-stratified distributions of mean of time-varying R(t) yields a median of 1.23 for women and 1.43 for men and a 95% CI of the difference as [-0.39, 0.07]. Comparing maximum of the time-varying R(t) yields a median value of 4.61 for women and 4.45 for men, with the 95% CI for their difference given by [-1.60, 1.65]. A group-wise comparison of most recent R(t) (as of June 3) yields a median of 0.79 for women and 1.05 for men, with 95% CI [-0.42, 0.09]. From *Figure 3*, we note that most women-led countries have median R(t) values towards the lower end of the scale. It appears that in terms of *<u>all features (barring the maximum) of the time varying reproduction number curve, women are ahead</u>*, but since all the confidence intervals contain zero, the results *are not statistically significant* at 5% level of significance.

Comparing the gender-stratified distribution of median of country-specific doubling times (longer is better for slowing down the virus), we note a similar pattern, with the distribution for women- countries having more mass on longer doubling times than men-led countries – with 16.6 and 16.1 days as the median values of doubling time for women and men respectively (the 95% bootstrap CI of the median difference of the two distributions is [-0.77, 3.17] days). The same goes for case-fatality rates (lower is better), women-led countries have lower median case-fatality rate, with the median for countries with women heads of state being 2.46%, while that for countries with men heads of state is 2.73%. Comparing CFRs stratified by gender of head of state by a two-sample bootstrap procedure yields a 95% CI of the median difference given by [-1.4%, 1.8%].

We know that extensive testing and contact tracing is key benchmark for success in this public health crisis. A comparison of percentage of population tested between the two groups yields median values of 3.28% and 1.59% for women- and men-led countries respectively (more testing is better). The 95% CI estimate of the difference of medians is [-1.29%, 3.60%]. Comparing women and men leaders on the basis of another testing metric – percentage of positive tests (lower is better) – yields a median 2.69% and 4.94% for women and men respectively, with [-4.89%, 0.30%] as the 95% CI of the difference of median values. *Figure 4* shows a visual overview of testing and we see women having on balance, more testing and lower test positives.

## Conclusion

Comparing three measures summarizing the trajectory of time varying reproduction number R(t) as well as most recent values of R(t), we note that the group of countries led by women appear to have better public health metrics measuring spread of the virus, although the median difference between the two groups of countries is not statistically significant. A similar comment can be made while investigating group-differences of doubling time and case-fatality rates. As far as scale of testing is concerned, we note that countries with women heads of state tend to do better with more testing and lower test positive rates, although this difference is again, not statistically significant. An interesting observation here is that all 18 countries with women heads of state included in our analysis are releasing testing data whereas only 67 out of the 141 countries led by men have testing data available.

This unadjusted ecological analysis is obviously wrinkled with several limitations and potential confounding that makes it impossible to conclude the causal impact of women leaders on pandemic outcomes. Several common causes can influence this pattern. Perhaps societies that are progressive and open enough to recruit diverse leaders and promote inclusive decision-making have a better chance of fighting the virus as a collective? Something to think about when we appoint and elect our future leaders.

## Data Availability

Data sourced from public repositories. All code are available at covind19.org. https://github.com/umich-cphds/cov-ind-19/blob/master/model/r_scripts/covind19_paper2.R

https://github.com/umich-cphds/cov-ind-19/blob/master/model/r_scripts/covind19_paper2.R

## Acknowledgments

The authors will like to thank the University of Michigan Rogel Cancer Center, Center for Precision Health Data Science, University of Michigan School of Public Health and Michigan Institute of Data Science for supporting this research:

## Data and materials availability

Data sourced from public repositories. All codes are available at covind19.org.

